# Why does COVID-19 case fatality rate vary among countries?

**DOI:** 10.1101/2020.04.17.20069393

**Authors:** Gabriele Sorci, Bruno Faivre, Serge Morand

## Abstract

**Background:** While the epidemic of SARS-CoV-2 is spreading worldwide, there is much concern over the mortality rate that the infection induces. Available data suggest that COVID-19 case fatality rate varies temporally (as the epidemic progresses) and spatially (among countries). Here, we attempted to identify key factors possibly explaining the variability in case fatality rate across countries.

**Methods:** We used data on the temporal trajectory of case fatality rate provided by the European Center for Disease Prevention and Control, and country-specific data on different metrics describing the incidence of known comorbidity factors associated with an increased risk of COVID-19 mortality at the individual level (Institute for Health Metrics and Evaluation). We also compiled data on demography, economy and political regimes for each country.

**Findings:** We first showed that temporal trajectories of case fatality rate greatly vary among countries. We found no evidence for association between comorbidities and case fatality rate at the country level. Case fatality rate was negatively associated with number of hospital beds x1,000 inhabitants. We also report evidence suggesting an association between case fatality rate and the political regime, with democracies suffering from the highest mortality burden, compared to autocratic regimes. However, most of the among-country variance in case fatality rate remained unexplained.

**Interpretation:** Overall, these results emphasize the role of socio-economic and political factors as possible drivers of COVID-19 case fatality rate at the country level.

**Funding:** None.

## Introduction

At the end of 2019, a novel coronavirus (SARS-CoV-2) emerged from an animal reservoir in the city of Wuhan, China^1,2^. Having established a human-to-human transmission, the virus rapidly spread, first within China, and subsequently outside China. Currently, more than 152 countries have reported at least one case of infection with SARS-CoV-2, and on 11^th^ March 2020 the WHO declared SARS-CoV-2 as a pandemic^3^. The coronavirus disease 2019 (COVID-19) produces a series of respiratory symptoms that vary in severity^4^. Although, in most infected people the infection results in mild symptoms that do not require particular medical care (or even in asymptomatic disease), in a substantial fraction of patients the disease develops into severe respiratory distress, requiring hospitalization in intensive care units^5^. In the absence of specific treatments that prevent or block viral replication, a serious matter of concern is the proportion of infected patients that will eventually die. Current data indicate that, worldwide, case fatality rate (CFR, the ratio between number of deaths and number of cases) might be around 2 – 3%. However, at the country level, CFR ranges from 0 to a frightening 12%^6^. There are many possible reasons for such a variation. First, the epidemic has spread in some countries earlier than in others and therefore the difference in CFR might reflect different stages during the spread of the disease. According to this hypothesis, one might expect an increase in CFR with time in countries where CFR is currently low. Second, at the individual level, clinical data have reported several risk factors associated with a poor prognosis. Age and comorbidities (cardiovascular diseases, cancers, diabetes mellitus, chronic lung diseases) seem to greatly increase the mortality risk^7-10^. Therefore, countries with a greater share of elderly in the population, or with higher incidence of recognized comorbidity factors might pay the highest toll to the infection. Third, as said before, in the absence of effective treatments, patients that develop severe symptoms require hospitalization in intensive care units for respiratory assistance. A major concern is that, as the number of infected people increases, the healthcare system will be overwhelmed^11^. Under this scenario, mortality rate might reflect the country-specific capacity to tackle a large number of patients requiring respiratory assistance and intensive care. Finally, countries might differ in their methodology to count and communicate the actual number of patients that have succumbed from SARS-CoV-2 infection.

Here, we conducted an analysis of the factors that might account for the variability of COVID-19 CFR among countries. Using data updated to 4^th^ Avril 2020, we first show that CFR significantly varies between countries, independently from the stage of the epidemic wave. Second, we used country-specific variables assessing the occurrence of comorbidities, as well as demographic, economic and political variables to uncover any association pattern with COVID-19 CFR. Overall, we could only account for a small fraction of the variance in CFR among countries, with no association between comorbidities and CFR. Among the demographic, economic and political variables, only number of hospital beds and the political regime were associated with CFR. These results emphasize the importance of strategic decisions for the management of health crisis during infectious disease pandemics.

## Methods

We used data on daily number of confirmed cases and deaths for each countries reported by the European Center for Disease Prevention and Control (ECDC). We computed the case fatality rate as the ratio between deaths and confirmed cases. We restricted the dataset to countries with at least 100 confirmed cases to avoid spurious results due to small numbers. For each country, we also counted the number of days between the 100^th^ case and 4^th^ Avril 2020, indicating the progression of the epidemic.

We used the online resource https://ourworldindata.org/ to retrieve data on the incidence of known comorbidity factors for each country, as well as information on demographics, economics and political regimes. In particular, we used different metrics to describe comorbidities: 1) disability-adjusted life years (DALYs); 2) age-standardized DALYs per 100,000; 3) share of total disease burden; 4) age-standardized death rates per 100,000; 5) share of deaths. We focused on known comorbidities such as cardiovascular diseases, cancers, chronic respiratory diseases, and diabetes. We also included metrics related to factors that might impinge on the severity of respiratory syndromes, such as smoking and air pollution. We also used https://ourworldindata.org/ to retrieve information on demographic, economic and political indicators. Table S1, in the supplementary information, reports the list, description and source of the variables used here, according to the GATHER statement^12^.

### Statistical analyses

We first aimed at exploring whether CFR varied among countries while controlling for the differences in the epidemic progression. To this purpose, we run a linear mixed model where CFR was the dependent variable, time since 100^th^ case (in days) and squared time since 100^th^ case (as to model non-linear variation), country and the two-way interactions between country and time were the fixed factors. Country was also included as a random effect. The covariance structure of R matrix was modeled using a first-order autoregression, to take into account the correlation between values from one day to the other. Degrees of freedom were computed using the Satterthwaite approximation. Only countries for which at least 10 days had elapsed between the record of the 100^th^ case and 4^th^ April 2020 were included in this model, to allow a better estimate of the variation between CFR and time. This model included 89 countries.

To assess the pattern of association between country-specific CFR (as assessed on 4^th^ Avril 2020) and comorbidities, we ran five multiple regression models that included different metrics. The first model included share of disease burden, the second model included share of deaths, the third model included DALYs per cause, the fourth model included age-standardized DALYs, and the fifth model included age-standardized death rates. All models always included a set of covariates that described demographic, economic and political variables, namely population size, life expectancy, share of the population over 70 years old, gross domestic product (GDP) per capita, total health care expenditure as share of GDP, number of hospital beds (x1,000 inhabitants), political regime. This last variable scores political regimes according to the level of democracy, between -10 (full autocracy) to +10 (full democracy) (Polity IV as reported in https://ourworldindata.org/). Finally, all models also included time to the 100^th^ case (in days), as to take into account the different progression of the epidemic in each country. Models were selected using a backward procedure with the criterion for staying in the model set at 0.05. For each variable included in the final model, we computed the partial R^2^. We also checked model fit and collinearity diagnostics (condition index^13^). All variables were standardized with mean = 0 and standard deviation = 1 to make parameter estimates directly comparable (variables with asymmetrical distribution were previously log-transformed to reduce skewness).

Different countries might have similar values of the different metrics used here because of their geographical proximity; in this case considering each country has a statistical unit might violate the assumption of independence of observations. We used Moran’s I coefficient to test if CFR and the explanatory variables were spatially autocorrelated (for each country we used the latitude and longitude of the capital to construct the distance matrix). We subsequently retested the association between CFR and explanatory variables using linear mixed models (LMMs) that, in addition to the fixed effects listed above (i.e., those included in the multiple regression models) also included three nested effects as random variables: continent (5 levels), geographic region (19 levels^14^) nested within continent, country (102 levels) nested within geographic region nested within continent.

To further explore the association between political regime and CFR, the -10 to 10 score was congregated into four classes indicating: 1) autocratic regimes (score from -10 to -6), 2) closed anocracy (from -5 to 0), 3) open anocracy (1 to 5), 4) democracy (6 to 10) (see https://ourworldindata.org/). Anocracy refers to political systems that combine features of both democratic and autocratic regimes, and that are therefore intermediate between the two^15^. This four level democracy score was included (as a fixed effect) in a LMM where CFR was the dependent variable. Time since the 100^th^ case and squared time plus the two-way interactions with the democracy score were also included as fixed effects, and the three nested effects accounting for the spatial clustering of countries were included as random effects. This model therefore informs us if countries with different levels of democracy have different trajectories of time-dependent CFR.

All the analyses were conducted with SAS 14.3.

## Results

We found strong evidence for among-country differences in COVID-19 CFR. The LMM showed a highly significant interaction between time since the 100^th^ case and country, indicating that CFR trajectories (both linear and quadratic components), do differ among countries as the epidemic progresses (Table 1, Figure 1).

**Table 1.** Linear mixed model exploring variation of COVID-19 case fatality rate (CFR) as a function of time since the 100^th^ case for each country. The model also included squared time since 100^th^ case and the interactions between country and time. Country was also declared as a random effect in the model. The model was restricted to countries that had 10 or more days elapsed between the occurrence of the 100^th^ case and 4^th^ April 2020 (to allow a better estimate of time-dependent variation in CFR). The analysis is based on 89 countries and 1,872 observations.

| *Fixed effects* | *F* | *df* | *p* |
| --- | --- | --- | --- |
| Country | 47.08 | 88,1605 | <0.0001 |
| Time since 100<sup>th</sup> case (days) | 5.72 | 1,1605 | 0.0169 |
| Squared time since 100<sup>th</sup> case | 11.47 | 1,1605 | 0.0007 |
| Country x time since 100<sup>th</sup> case | 13.80 | 88,1605 | <0.0001 |
| Country x squared time since 100<sup>th</sup> case | 11.40 | 88,1605 | <0.0001 |

**Figure 1.**
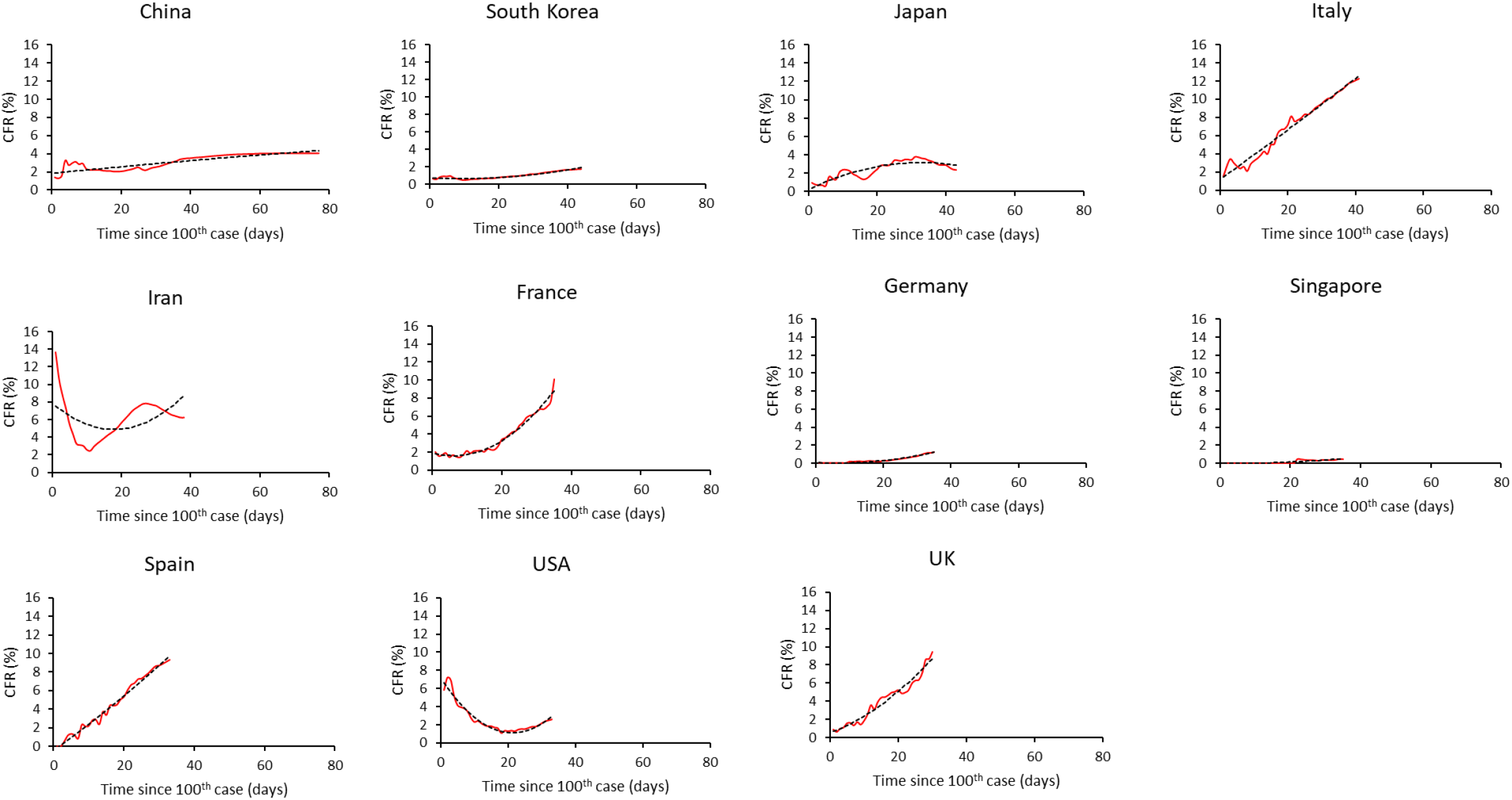
Time-dependent variation in COVID-19 case fatality rate (CFR). Time refers to the number of days between the occurrence of the 100^th^ case and 4th April 2020. The different panels refer to countries with at least 30 days elapsed since the occurrence of the 100^th^ case. Red lines describe the observed values, the dotted lines refer to the predictions of a linear mixed model.

In order to uncover the factors that might account for such among-country variation in CFR, we ran five multiple regression models that included different explanatory variables related to comorbidities, demographic, economic and political factors. These models used CFR estimated on 4^th^ April 2020 as the dependent variable. Among the comorbidity metrics, none was associated with CFR, the only exception being the model on DALYs that showed a positive association between DALYs lost to diabetes, urogenital, blood and endocrine diseases and CFR (Table 2). It should be noted that DALYs are highly correlated with population size and therefore, it is possible that this is a spurious result due to the covariation between the two variables. Using other metrics of comorbidities (share of disease burden, share of deaths, age-standardized death rates ×100,000, age-standardized DALYs ×100,000) produced no association with CFR (Table 2). Four demographic, economic and political variables were associated with CFR across different models (Table 2). Population size was positively associated with CFR in all models, except the one on DALYs, corroborating the idea that the above reported association between DALYs lost to diabetes, urogenital, blood and endocrine diseases might merely result from the covariation with population size. Number of hospital beds per 1,000 inhabitants was negatively associated with CFR in all models run. Life expectancy (in four models) and share of the population over 70 years old (in one model) were positively associated to CFR. Finally, in four models, we found a positive association between CFR and level of democracy. Overall, the amount of CFR variance accounted by the different factors (comorbidities, demographics, economics, politics) was small (around 20% of variance explained by the model), and the partial amount of variance explained by the factors that were retained in the final models was very small (ranging from 2% to 8%).

**Table 2.** Multiple regression models investigating the association between COVID-19 case fatality rate (CFR) and several descriptors of comorbidities, demographics, economics and political regime for each country (n = 102). Each model included the same demographic, economic and political regime variables (GDP per capita, population size, total health care expenditure as share of GDP, number of hospital beds x1,000 inhabitants, share of the population over 70 years, political regime). In addition, model 1 included variables referring to the share of total disease burden due to cardiovascular diseases, cancers, chronic respiratory diseases, diabetes. Model 2 included variables referring to share of deaths for cardiovascular diseases, cancers, chronic respiratory diseases, lower respiratory diseases, diabetes, outdoor air pollution. Model 3 included variables referring to DALYs lost to cardiovascular diseases, cancers, chronic respiratory diseases, diabetes. Model 4 included variables referring to age-standardized DALYs lost per 100,000 people. Model 5 included variables referring to age-standardized death rates due to cardiovascular diseases, cancers, smoking, air pollution, ambient particulate air pollution, pneumonia over 70 years, outdoor air pollution over 70 years, smoking over 70 years. The table reports the variables selected by the backward procedure of the multiple regression. Models 1, 2, and 4 retained the same variables in the final model, therefore only the results of model 1 are reported in the table. We report model R^2^ and p-values, as well as the standardized slope (95% CI), partial R^2^ and p-values of all variables retained in the final models.

| *Model 1 (share of total disease burden) $R^2 = 0.195$, $p = 0.0003$* | *standardized slope (95% CI)* | *t value* | *partial $R^2$* | *p* |
| --- | --- | --- | --- | --- |
| Population size | 0.910<br>(0.342/1.477) | 3.18 | 0.080 | 0.0020 |
| Political regime | 0.729<br>(0.212/1.246) | 2.80 | 0.065 | 0.0062 |
| Number of hospital beds | -0.691<br>(-1.278/-0.105) | -2.34 | 0.028 | 0.0214 |
| Life expectancy | 0.633<br>(0.053/1.214) | 2.16 | 0.022 | 0.0329 |
| *Model 3 (DALYs per cause) $R^2 = 0.203$, $p = 0.0002$* | *standardized slope (95% CI)* | *t value* | *partial $R^2$* | *p* |
| DALYs lost to diabetes | 0.967<br>(0.392/1.543) | 3.33 | 0.086 | 0.0012 |
| Political regime | 0.720<br>(0.205/1.235) | 2.78 | 0.063 | 0.0066 |
| Life expectancy | 0.678<br>(0.095/1.259) | 2.31 | 0.028 | 0.0230 |
| Number of hospital beds | -0.668<br>(-1.253/-0.082) | -2.26 | 0.025 | 0.0258 |
| *Model 5 (age-standardized death rates) $R^2 = 0.178$, $p = 0.0002$* | *standardized slope (95% CI)* | *t value* | *partial $R^2$* | *p* |
| Population size | 0.742<br>(0.179/1.306) | 2.61 | 0.080 | 0.0103 |
| Number of hospital beds | -1.037<br>(-1.749/-0.325) | -2.89 | 0.070 | 0.0047 |
| Share of the population over 70 | 1.182<br>(0.447/1.889) | 3.31 | 0.027 | 0.0013 |

Neighboring countries can have similar values of CFR, comorbidities, economic, demographic and political variables, because of their geographical proximity, violating the assumption of independence of statistical units. A spatial autocorrelation analysis showed, indeed, positive autocorrelation for CFR, life expectancy, share of the population over 70 years, number of hospital beds, and political regime (Moran’s I, all p’s <0.0001). We therefore reran the association models using five LMMs that incorporated the spatial clustering of countries (into continents and geographical regions). These LMMs confirmed 1) the lack of association between CFR and comorbidity factors, 2) the negative association between number of hospital beds and CFR, 3) the positive association between level of democracy and CFR (Table 3). Contrary to the regression models, LMMs that integrated spatial information did not provide evidence for an association between life expectancy (or share of the population over 70 years) and CFR (Table 3).

**Table 3.** Linear mixed effects models exploring the associations between COVID-19 case fatality rate (CFR) and different comorbidity, demographic, economic, and political variables. The models included three nested factors (continent, geographical region and country) as random effects. The table reports parameter estimates (with SE and 95% CI), t and p values. Sample size is five continents, 19 geographical regions, 102 countries and 1,879 observations.

| *Model 1 (share of total disease burden)* |  |  |  |  |  |
| --- | --- | --- | --- | --- | --- |
| *Fixed effects* | Estimate | SE | t | p | 95% CI |
| Intercept | 1.798 | 0.939 |  |  |  |
| Time since 100<sup>th</sup> case | 1.168 | 0.062 | 18.93 | <0.0001 | 1.047/1.289 |
| Squared time since 100<sup>th</sup> case | -0.393 | 0.063 | -6.27 | <0.0001 | -0.516/-0.270 |
| Cardiovascular | 0.206 | 0.271 | 0.76 | 0.4501 | -0.333/0.744 |
| Cancer | -0.496 | 0.552 | -0.90 | 0.3710 | -1.595/0.602 |
| Diabetes | 0.360 | 0.226 | 1.59 | 0.1143 | -0.089/0.809 |
| Chronic respiratory | -0.108 | 0.261 | -0.41 | 0.681 | -0.626/0.410 |
| Population size | 0.717 | 0.297 | 2.41 | 0.018 | 0.126/1.308 |
| GDP per capita | -0.538 | 0.294 | -1.83 | 0.0704 | -1.122/0.046 |
| Life expectancy | 0.194 | 0.365 | 0.53 | 0.5968 | -0.533/0.921 |
| Share of the population over 70 | 0.160 | 0.582 | 0.28 | 0.7840 | -0.999/1.320 |
| Number of hospital beds | -0.923 | 0.347 | -2.66 | 0.0094 | -1.613/-0.233 |
| Total health care expenditure (share of GDP) | -0.189 | 0.283 | -0.67 | 0.508 | -0.753/0.376 |
| Level of democracy | 0.650 | 0.292 | 2.23 | 0.0285 | 0.070/1.229 |
| *Covariance parameters* | Estimate | SE | z | p |  |
| Variance | 2.414 | 0.384 | 6.29 | <0.0001 |  |
| First-order autoregression | 0.127 | 0.144 | 0.88 | 0.3781 |  |
| Residual | 1.089 | 0.037 | 29.77 | <0.0001 |  |
| *Model 2 (share of deaths)* |  |  |  |  |  |
| *Fixed effects* | Estimate | SE | t | p | 95% CI |
| Intercept | 1.823 | 0.953 |  |  |  |
| Time since 100<sup>th</sup> case | 1.169 | 0.062 | 18.93 | <0.0001 | 1.047/1.290 |
| Squared time since 100<sup>th</sup> case | -0.393 | 0.063 | -6.27 | <0.0001 | -0.516/-0.270 |
| Cardiovascular | 0.054 | 0.373 | 0.14 | 0.8858 | -0.689/0.796 |
| Cancer | -0.676 | 0.519 | -1.30 | 0.1967 | -1.710/0.357 |
| Chronic respiratory | -0.165 | 0.249 | -0.66 | 0.5105 | -0.660/0.331 |
| Lower respiratory | -0.130 | 0.263 | -0.49 | 0.624 | -0.653/0.394 |
| Diabetes | 0.202 | 0.231 | 0.88 | 0.3832 | -0.257/0.661 |
| Outdoor air pollution | -0.023 | 0.322 | -0.07 | 0.9438 | -0.663/0.617 |
| Population size | 0.669 | 0.309 | 2.17 | 0.0331 | 0.055/1.284 |
| GDP per capita | -0.383 | 0.307 | -1.25 | 0.2166 | -0.994/0.229 |
| Life expectancy | 0.412 | 0.395 | 1.04 | 0.3002 | -0.374/1.197 |
| Share of the population over 70 | 0.202 | 0.506 | 0.40 | 0.6909 | -0.805/1.210 |
| Number of hospital beds | -0.961 | 0.351 | -2.74 | 0.0075 | -1.658/-0.264 |
| Total health care expenditure<br>(share of GDP) | -0.195 | 0.283 | -0.69 | 0.4947 | -0.759/0.370 |
| Level of democracy | 0.638 | 0.305 | 2.09 | 0.0395 | 0.031/1.245 |
| *Covariance parameters* | Estimate | SE | z | p |  |
| Variance | 2.462 | 0.396 | 6.22 | <0.0001 |  |
| First-order autoregression | 0.135 | 0.135 | 1.00 | 0.318 |  |
| Residual | 1.089 | 0.037 | 29.76 | <0.0001 |  |
| *Model 3 (DALYs lost)* |  |  |  |  |  |
| *Fixed effects* | Estimate | SE | t | p | 95% CI |
| Intercept | 1.813 | 0.900 |  |  |  |
| Time since 100<sup>th</sup> case | 1.168 | 0.062 | 18.93 | <0.0001 | 1.047/1.289 |
| Squared time since 100<sup>th</sup> case | -0.393 | 0.063 | -6.27 | <0.0001 | -0.516/-0.270 |
| Cardiovascular | 1.805 | 1.253 | 1.44 | 0.1534 | -0.686/4.2961 |
| Cancer | -3.473 | 1.962 | -1.77 | 0.0805 | -7.377/0.432 |
| Chronic respiratory | -0.391 | 1.609 | -0.24 | 0.8086 | -3.589/2.807 |
| Diabetes | 2.476 | 1.293 | 1.91 | 0.0587 | -0.093/5.045 |
| Population size | 0.312 | 1.942 | 0.16 | 0.8728 | -3.548/4.172 |
| GDP per capita | -0.538 | 0.288 | -1.87 | 0.0651 | -1.110/0.034 |
| Life expectancy | 0.388 | 0.358 | 1.08 | 0.2815 | -0.324/1.100 |
| Share of the population over 70 | 0.337 | 0.580 | 0.58 | 0.5625 | -0.817/1.492 |
| Number of hospital beds | -0.968 | 0.340 | -2.85 | 0.0055 | -1.643/-0.293 |
| Total health care expenditure<br>(share of GDP) | -0.222 | 0.275 | -0.81 | 0.4217 | -0.768/0.325 |
| Level of democracy | 0.657 | 0.284 | 2.31 | 0.0230 | 0.093/1.221 |
| *Covariance parameters* | Estimate | SE | z | p |  |
| Variance | 2.287 | 0.361 | 6.34 | <0.0001 |  |
| First-order autoregression | 0.097 | 0.134 | 0.72 | 0.4702 |  |
| Residual | 1.089 | 0.037 | 29.76 | <0.0001 |  |

| *Model 4 (DALYs lost x100,000)* |  |  |  |  |  |
| --- | --- | --- | --- | --- | --- |
| *Fixed effects* | Estimate | SE | t | *p* | 95% CI |
| Intercept | 1.788 | 0.956 |  |  |  |
| Time since 100<sup>th</sup> case | 1.168 | 0.062 | 18.93 | <0.0001 | 1.047/1.289 |
| Squared time since 100<sup>th</sup> case | -0.393 | 0.063 | -6.27 | <0.0001 | -0.516/-0.270 |
| DALYs lost all ages | -0.173 | 0.610 | -0.28 | 0.7770 | -1.385/1.039 |
| DALYs lost over 70 | -0.459 | 0.435 | -1.06 | 0.2943 | -1.324/0.406 |
| DALYs lost age-standardized | 1.204 | 1.096 | 1.10 | 0.2750 | -0.975/3.382 |
| Population size | 0.420 | 0.276 | 1.52 | 0.1318 | -0.129/0.969 |
| GDP per capita | -0.468 | 0.319 | -1.47 | 0.1459 | -1.103/0.166 |
| Life expectancy | 0.527 | 0.537 | 0.98 | 0.3290 | -0.541/1.595 |
| Share of the population over 70 | 0.295 | 0.600 | 0.49 | 0.6244 | -0.898/1.488 |
| Number of hospital beds | -0.868 | 0.349 | -2.49 | 0.0148 | -1.563/-0.174 |
| Total health care expenditure (share of GDP) | -0.326 | 0.280 | -1.16 | 0.2476 | -0.882/0.231 |
| Level of democracy | 0.481 | 0.269 | 1.79 | 0.0768 | -0.053/1.014 |
| *Covariance parameters* | Estimate | SE | z | *p* |  |
| Variance | 2.483 | 0.393 | 6.31 | <0.0001 |  |
| First-order autoregression | 0.133 | 0.142 | 0.94 | 0.3471 |  |
| Residual | 1.089 | 0.037 | 29.76 | <0.0001 |  |

| *Model 5 (age-standardized death rates x100,000)* |  |  |  |  |  |
| --- | --- | --- | --- | --- | --- |
| *Fixed effects* | Estimate | SE | t | *p* | 95% CI |
| Intercept | 1.822 | 0.912 |  |  |  |
| Time since 100<sup>th</sup> case | 1.169 | 0.062 | 18.94 | <0.0001 | 1.048/1.290 |
| Squared time since 100<sup>th</sup> case | -0.394 | 0.063 | -6.28 | <0.0001 | -0.517/-0.271 |
| Cardiovascular | 0.351 | 0.507 | 0.69 | 0.4908 | -0.660/1.360 |
| Cancer | -0.372 | 0.246 | -1.51 | 0.1346 | -0.862/0.118 |
| Smoking | -0.333 | 0.653 | -0.51 | 0.6111 | -1.632/0.965 |
| Air pollution | 0.405 | 0.911 | 0.44 | 0.658 | -1.407/2.218 |
| Ambient particulate air pollution | 1.217 | 1.541 | 0.79 | 0.4323 | -1.850/4.283 |
| Pneumonia over 70 | -0.160 | 0.265 | -0.60 | 0.5474 | -0.686/0.367 |
| Outdoor air pollution over 70 | -1.113 | 1.254 | -0.89 | 0.3774 | -3.609/1.383 |
| Smoking over 70 | 0.233 | 0.488 | 0.48 | 0.6343 | -0.738/1.203 |
| Population size | 0.679 | 0.282 | 2.41 | 0.0181 | 0.119/1.239 |
| GDP per capita | -0.271 | 0.389 | -0.70 | 0.4883 | -1.046/0.504 |
| Life expectancy | 0.358 | 0.405 | 0.89 | 0.3784 | -0.446/1.163 |
| Share of the population over 70 | 0.256 | 0.499 | 0.51 | 0.6099 | -0.739/1.250 |
| Number of hospital beds | -0.973 | 0.357 | -2.73 | 0.0077 | -1.683/-0.264 |
| Total health care expenditure<br>(share of GDP) | -0.109 | 0.298 | -0.37 | 0.7137 | -0.701/0.482 |
| Level of democracy | 0.690 | 0.297 | 2.32 | 0.0226 | 0.099/1.280 |
| *Covariance parameters* | Estimate | SE | z | p |  |
| Variance | 2.447 | 0.395 | 6.19 | <0.0001 |  |
| First-order autoregression | 0.081 | 0.155 | 0.52 | 0.560 |  |
| Residual | 1.089 | 0.037 | 29.76 | <0.0001 |  |

To further explore the association between democracy level and time-dependent variation in CFR, we grouped countries as having an autocratic, closed anocratic, open anocratic or democratic political regime. We then ran a LMM to test whether time-dependent trajectories of CFR differed according to this four level variable. The LMM also included the information on spatial clustering (nested factors declared as random effects). The model showed highly significant interactions between time since 100^th^ case (both linear and quadratic components) and political regime (Table 4, Figure 2). Therefore, CFR increased with a steeper slope in countries with democratic regimes compared to less democratic political systems (Figure 2).

**Table 4.** Linear mixed effect models exploring the association between political system (autocracy, closed anocracy, open anocracy, democracy) and CFR. The model included time since 100^th^ case and squared time and the interactions between time and political system. The model also included three nested factors (continent, geographical region and country) as random effects. Sample size is five continents, 18 geographical regions, 103 countries and 1,981 observations.

| *Fixed effects* | *F* | *df* | *p* |
| --- | --- | --- | --- |
| Political system | 0.65 | 3,108 | 0.5823 |
| Time since 100<sup>th</sup> case (days) | 14.39 | 1,1877 | 0.0002 |
| Squared time since 100<sup>th</sup> case | 0.71 | 1,1876 | 0.4007 |
| Political system x time since 100<sup>th</sup> case | 6.87 | 3,1879 | 0.0001 |
| Political system x squared time since 100<sup>th</sup> case | 12.44 | 3,1879 | <0.0001 |

**Figure 2.**
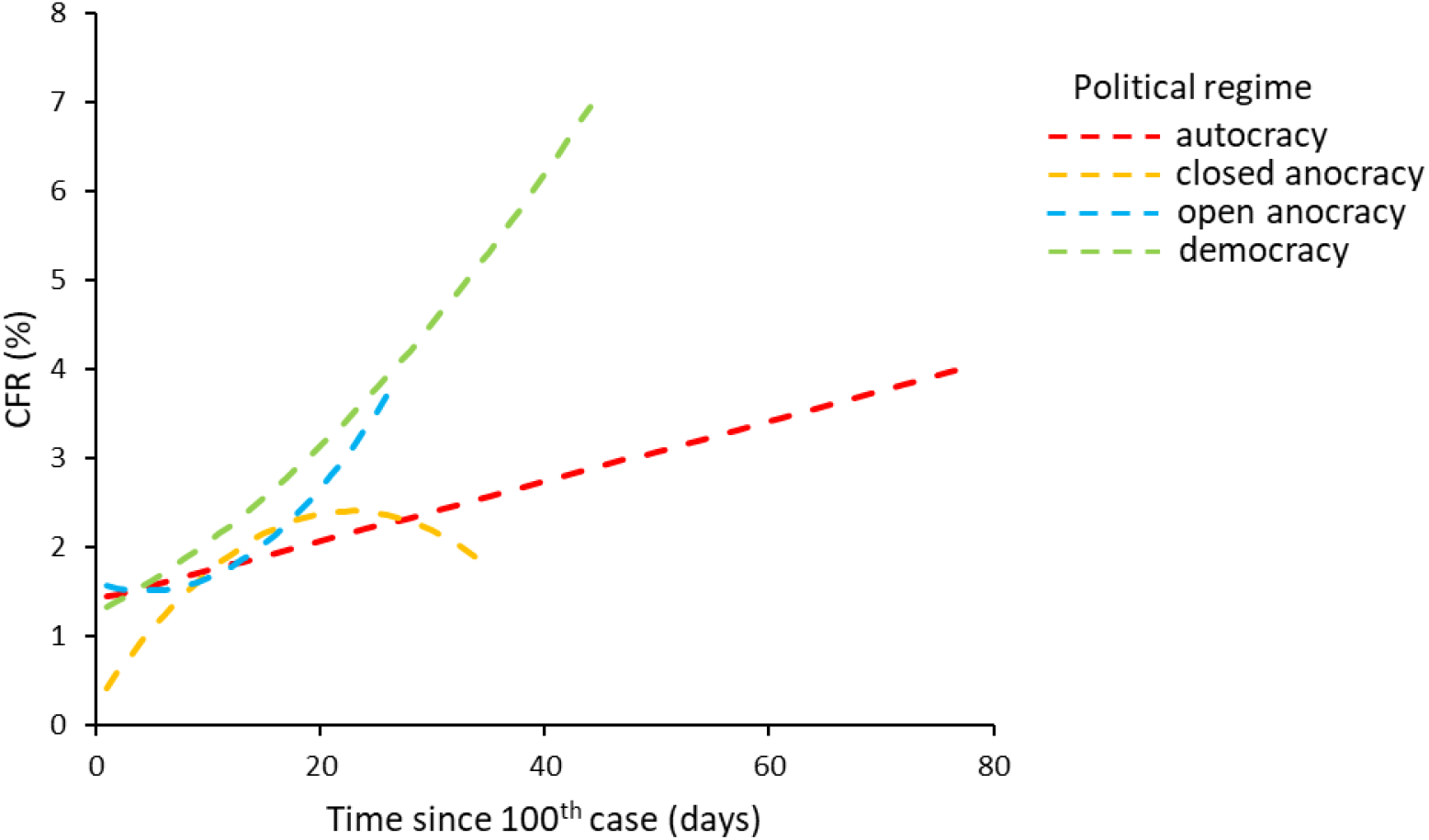
Time-dependent variation in COVID-19 case fatality rate (CFR) according to the political regime. Time refers to the number of days between the occurrence of the 100^th^ case and 4^th^ April 2020. The different lines refer to the predictions of a linear mixed model (red = autocracy, orange = closed anocracy, blue = open anocracy, green = democracy).

## Discussion

As the SARS-CoV-2 epidemic continues to spread worldwide, there is an increasing concern related to the mortality induced by the disease. Currently, some European countries are paying the highest toll to the infection, with case fatality rate between 10 and 12%. In this light, it is important to understand why some countries seem to experience lower mortality rate than others and possibly to predict how CFR might change as the epidemic progresses.

Assessing case fatality rate during an ongoing outbreak is particularly difficult for, at least two reasons^6,16^. CFR at a given time might be an underestimate of the final mortality rate because cumulative number of deaths will eventually keep increasing as some patients are still hosted in intensive care units. Conversely, CFR might overestimate actual mortality if a large fraction of infected people do not develop disease symptoms (or develop mild symptoms) and therefore this fraction is not included into the population of confirmed infection cases.

Despite the uncertainty associated with the estimation of CFR and how it will evolve over time, its comparison between countries can provide useful insights into the heterogeneity in the current burden paid to the disease. We showed that the trajectories of time dependent variation in CFR greatly differed between countries. This quantitatively corroborates and statistically validates the intuition that some countries are better dealing with the disease than others.

The following step was to try to understand whether such heterogeneity arises as the consequence of predictable factors. Previous reports on the clinical outcome of the disease have identified two major factors associated with poor prognosis: age and the presence of comorbidities. Elderly people have been shown to suffer the highest mortality rate following infection with SARS-CoV-2, with case fatality rates possibly higher than 20% in patients older than 80 years^6,17^. Similarly, previous history of cardiovascular disorders, cancer and diabetes have been reported to substantially increase COVID-19 mortality risk^5,18^. We therefore predicted that countries with a higher share of elderly people and a higher incidence of known comorbidity factors might suffer from the highest CFR. However, the results did not provide any support for this prediction. Life expectancy and share of populations over 70 years were actually positively associated with CFR when running multiple regression models, but these associations did not receive statistical support when using models that explicitly took into account the spatial clustering of countries.

Economic and political parameters might equally well contribute to shape COVID-19 mortality. As the number of severe cases increases during the epidemic, the health care system can get overwhelmed and might be unable to receive and treat all those who need intensive care. Mortality might therefore results from health care systems that are inadequate to deal with large number of cases requiring simultaneous admittance in intensive care units. We used several proxies describing the investment of each country into the health care system and found a negative association between the number of hospital beds per 1,000 inhabitants and CFR, and this result was robust to the inclusion of spatial clustering into the model. This result is therefore in agreement with the idea that socio-economic factors are currently playing a role to shape COVID-19 CFR.

A final source of variation accounting for differences in CFR might be due to differential reports of number of deaths and/or confirmed cases between countries. This might reflect different counting/reporting methodology (e.g., deciding whether or not a given patient died because of COVID-19). In addition, implementing effective social distancing protocols, that are currently deployed in most of the countries facing the epidemic, might prove more difficult in democratic countries, compared to other political regimes that exert a more stringent control over the population. In agreement with this hypothesis, we found that countries with a democratic political regime had the highest CFR. A further model showed that time-dependent trajectories also varied among countries according to their political regimes, with democracies having the steepest increase of CFR as the epidemic progresses. The role of social and cultural traits in the emergence of zoonotic diseases has already been discussed in the past, including the idea that collectivistic societies might have built as a way to better control epidemic waves^19^. In agreement with this hypothesis, Morand and Walther^20^ found that collectivistic societies were less exposed to zoonotic disease outbreaks compared to individualistic ones.

Although we report here some associations between economic, political variables and COVID-19 CFR, we fully acknowledge that these factors explain a little fraction of the total variation in CFR among countries. This might come from the coarse grain of the analyses (country level), the error associated with the metrics used in our study, or the uncertainties associated with the estimation of CFR while the epidemic is still ongoing. When serological tests will be available, a large-scale assessment of the proportion of the population that has been infected by the virus will provide more accurate data to better estimate COVID-19 CFR and the possible factors explaining the among-country heterogeneity. With this in mind, we nevertheless believe that our results stress the role of socio-economic and political factors as potential drivers affecting how a country deals with globally threatening epidemics. In particular, past decisions on the policy of investment into the healthcare system have the potential to severely impact the whole society when a major sanitary crisis outburst. Another very challenging issue that many countries are currently facing, at the crossroads of our modern lifestyle, is to extent to which democracies are willing to renounce to individual freedom for a collectivistic benefit^21^.

## Data Availability

All data used in this manuscript come from open online resources

## Contributors

GS, BF and SM conceived the study; GS compiled the data and analyzed them; GS, BF, and SM interpreted the results; GS wrote the first draft of the manuscript; GS, BF and SM revised the manuscript and approved the final version.

## References

1. Zhu N, Zhang D, Wang W, et al. A Novel Coronavirus from Patients with Pneumonia in China, 2019. N Engl J Med 2020; 382: 727–733, doi.org/10.1056/NEJMoa2001017.

2. Zhou P, Yang X-L, Wang X-G, et al. A pneumonia outbreak associated with a new coronavirus of probable bat origin. Nature 2020; published online 3 February 2020, doi.org/10.1038/s41586-020-2012-7.

3. WHO Director-General’s opening remarks at the media briefing on COVID-19 - 11 March 2020. https://www.who.int/dg/speeches/detail/who-director-general-s-opening-remarks-at-the-media-briefing-on-covid-1911-march-2020.

4. Wu Z, McGoogan JM. Characteristics of and important lessons from the coronavirus disease 2019 (COVID-19) outbreak in China. Summary of a report of 72314 cases from the Chinese center for disease control and prevention. JAMA 2020; published online 24 February 2020, doi.org/10.1001/jama.2020.2648.

5. Wang D, Hu B, Hu C, et al. Clinical characteristics of 138 hospitalized patients with 2019 novel coronavirus-infected pneumonia in Wuhan, China. JAMA 2020; 323: 1061–1069, doi.org/10.1001/jama.2020.1585.

6. Kim D-H, Choe YJ, Jeong J-Y. Understanding and interpretation of case fatality rate of coronavirus disease 2019. J Korean Med Sci 2020 ; 30: e137, doi.org/10.3346/jkms.202035.e137.

7. CDC COVID-19 Response Team. Preliminary Estimates of the Prevalence of Selected Underlying Health Conditions Among Patients with Coronavirus Disease 2019 — United States, February 12–March 28, 2020. MMWR Morb Mortal Wkly Rep 2020; 69: 382–386. doi.org/10.15585/mmwr.mm6913e2.

8. Chen N, Zhou M, Dong X et al. Epidemiological and clinical characteristics of 99 cases of 2019 novel coronavirus pneumonia in Wuhan, China: a descriptive study. Lancet 2020; 395: 507–513, doi.org/10.1016/S0140-6736(20)30211-7.

9. Huang C, Wang Y, Li X, et al. Clinical features of patients infected with 2019 novel coronavirus in Wuhan, China. Lancet 2020; 395: 497–506, doi.org/10.1016/S0140-6736(20)30183-5.

10. Lescure F-X, Bouadma L, Nguyen D, Clinical and virological data of the first cases of COVID-19 in Europe: a case series. Lancet Infect Dis 2020; published online 27 March 2020, doi.org/10.1016/S1473-3099(20)30200-0.

11. Ji Y, Ma Z, Peppelenbosch MP, Pan Q. Potential association between COVID-19 mortality and health-care resource availability. Lancet Glob Health 2020; published online 25 February 2020, doi.org/10.1016/S2214-109X(20)30068-1.

12. Stevens GA, Alkema L, Black RE, et al. Guidelines for Accurate and Transparent Health Estimates Reporting: the GATHER statement. Lancet 2016; 388: e19–23, doi.org/10.1016/S0140-6736(16)30388-9.

13. Belsley DA, Kuh E, Welsch RE,Regression Diagnostics: Identifying Influential Data and Sources of Collinearity. John Wiley & Sons, New York (1980).

14. United Nations. Standard country or area codes for statistical use (M49). 2017. https://unstats.un.org/unsd/methodology/m49/.

15. Epstein D, Bates R, Goldstone J, Kristensen I. Democratic Transitions. Am J Pol Sci 2006; 50: 551–568.

16. Rajgor DD, Lee MH, Archuleta S, Bagdasarian N, Quek SC. The many estimates of the COVID-19 case fatality rate. Lancet Infect Dis 2020; published online 27 March 2020, doi.org/10.1016/S1473-3099(20)30244-9.

17. Ruan Q, Yang K, Wang W, Jiang L, Song J. Clinical predictors of mortality due to COVID-19 based on an analysis of data of 150 patients from Wuhan, China. Intensive Care Med 2020; published online 3 March 2020, doi.org/10.1007/s00134-020-05991-x.

18. Yang X, Yu Y, Xu J, et al. Clinical course and outcomes of critically ill patients with SARS-CoV-2 pneumonia in Wuhan, China: a single-centered, restrospective, observational study. Lancet Respir Med, published online 21 February 2020, doi.org/10.1016/S2213-2600(20)30079-5.

19. Fincher CL, Thornhill R, Murray DR, Schaller M. Pathogen prevalence predicts human cross-cultural variability in individualism/collectivism. Proc R Soc Lond B 2008; 275: 1279–1285.

20. Morand S, Walther B. Individualistic values are related to an increase in the outbreaks of infectious diseases and zoonotic diseases. Sci Rep 2018; 8: 3866, doi.org/10.1038/s41598-018-22014-4.

21. Studdert DM, Hall MA. Disease Control, Civil Liberties, and Mass Testing — Calibrating Restrictions during the Covid-19 Pandemic. N Engl J Med 2020; published online 9 April 2020, doi.org/10.1056/NEJMp2007637.

